# Seroprevalence and immunity of SARS-CoV-2 infection in children and adolescents in schools in Switzerland: design for a longitudinal, school-based prospective cohort study

**DOI:** 10.1101/2020.08.30.20184671

**Authors:** Agne Ulyte, Thomas Radtke, Irène Abela, Sarah R Haile, Julia Braun, Ruedi Jung, Christoph Berger, Alexandra Trkola, Jan Fehr, Milo A. Puhan, Susi Kriemler

## Abstract

**Introduction:** Seroprevalence and transmission routes of severe acute respiratory syndrome coronavirus 2 (SARS-CoV-2) infection in children and adolescents, especially in school setting, are not clear. Resulting uncertainty is reflected in very different decisions on school closures and reopenings across countries. The aim of this longitudinal cohort study is to assess the extent and patterns of seroprevalence of SARS-CoV-2 antibodies in school-attending children repeatedly. It will examine risk factors for infection, relationship between seropositivity and symptoms, and temporal persistence of antibodies. Additionally, it will include testing of school personnel and parents.

**Methods and analysis:** The study *(Ciao Corona)* will enroll a regionally representative, random sample of schools in the canton of Zurich, where 18% of the Swiss population live. Children aged 5 to 16 years, attending classes in primary and secondary schools are invited. Venous blood and saliva samples are collected for SARS-CoV-2 serological testing after the first wave of infections (June/July 2020), in fall (October/November 2020), and after winter (March/April 2021). Venous blood is also collected for serological testing of parents and school personnel. Bi-monthly questionnaires to children, parents and school personnel cover SARS-CoV-2 symptoms and tests, health, preventive behavior, lifestyle and quality of life information. Total seroprevalence and cumulative incidence will be calculated. Hierarchical Bayesian logistic regression models will account for sensitivity and specificity of the serological test in the analyses and for the complex sampling structure, i.e., clustering within classes and schools.

**Ethics and dissemination:** The study was approved by the Ethics Committee of the Canton of Zurich, Switzerland (2020-01336). The results of this study will be published in peer-reviewed journals and will be made available to study participants and participating schools, the Federal Office of Public Health, and the Educational Department of the canton of Zurich. **Trial registration number** NCT04448717.

- *Ciao Corona* is a large, prospective school-based cohort study and will provide robust data on severe acute respiratory syndrome coronavirus 2 (SARS-CoV-2) seroprevalence, transmission routes and immunity over time in a representative sample of school children.
- The longitudinal design will allow describing temporal trends of immunity to SARS-CoV-2 infection and to evaluate effects of school structure and preventive measures.
- This study will inform goal-oriented policy decisions in school management during subsequent outbreaks.
- Participation bias, missing questionnaires, desirability bias, and loss of follow up may occur. The validity of serological tests may also hamper results.

## Introduction

Decisions on school opening or closing during the severe acute respiratory syndrome coronavirus 2 (SARS-CoV-2) pandemic vary greatly across and even within countries. While some countries kept schools mostly open (e.g., Sweden, Australia) or reopened early (e.g., Denmark), others opted for prolonged closing with decision on reopening in late summer 2020 pending (e.g., the US, Italy, Ireland). Early school closure in response to pandemic was partly guided by analogy of transmission of other viruses, such as influenza [1,2], but current reports suggest that the susceptibility and transmissibility of children may be largely different for SARS-CoV-2 compared [3]. Lower prevalence of SARS-CoV-2 infection is reported in younger children – potentially as they are infected less frequently or as infections are more frequently asymptomatic and thus underestimated [4–6]. Although children may infect others less often than adults, their exact role in transmission pathways is still not clear [7].

A few studies have reported population-level seroprevalence of SARS-CoV-2, including the population of children [4,5]. Such studies focused on households, leaving the role of schools in SARS-CoV-2 transmission unclear, especially as they were often conducted during school closure. As most of school-aged children and adolescents’ social interactions take place in family and school [8], schools could play a crucial role in spreading the infection. Currently, mostly anecdotal evidence and case studies of SARS-CoV-2 infection spread in schools exist [9,10].

Many schools were or still are closed worldwide in response to the pandemic, without solid arguments for or sufficient understanding of potential consequences of such a policy [11]. 1.6 billion learners worldwide (90% of all) were affected by school closure [12]. In the US, school closure was coordinated on state and district level, with all US public schools eventually closing from March 25^th^, 2020 [13]. The opening of the schools might be delayed in many districts, and the exact mode of teaching is still not clear [14]. In Switzerland, schools were closed from March 16^th^ to May 10^th^, 2020 (switching to home and online schooling), then partly reopened until June 7^th^ (e.g., teaching in half-classes, restricting larger group activities, reducing school care groups), when regular teaching resumed again [15].

There is an urgent need for representative, population-based studies on children and adolescents especially in the school setting, to answer questions about the prevalence, infection routes, asymptomatic cases, risk factors, and duration of immunity to SARS-CoV-2 infection. This article reports the design and protocol of a longitudinal school-based seroprevalence study conducted in the largest canton of Switzerland. The study is part of a large nationally coordinated research network *Corona Immunitas*, and one of the first and largest representative studies of SARS-CoV-2 spread in children and adolescents in schools, globally.

## Methods and analysis

### Study overview, design and population

#### Study objectives

The study focuses on the seroprevalence and potential clustering of SARS-CoV-2 infection in children and adolescents attending school, as well as history, symptoms, and risk factors for SARS-CoV-2, health, lifestyle and quality of life outcomes. It aims to address the following objectives:

1. To repeatedly determine the seroprevalence of SARS-CoV-2 antibodies in school-aged children covering grades one to eight (approximately 6-16 years old) after the lockdown and the subsequent reopening of schools (June/July 2020), three months after the start of the next school year (October/November 2020), and after the winter (March/April 2021);
2. To examine clustering of seropositive cases within classes, schools and districts, and temporal evolution of the clusters;
3. To determine the proportion of asymptomatic children and adolescents with SARS-CoV-2 antibodies;
4. To determine the duration of the acquired immunity by examining new infections in children with positive serology and temporal persistence of detectable SARS-CoV-2 antibodies;
5. To identify sociodemographic, exposure, hygiene, school- and family-based behavioral and environmental risk factors for SARS-CoV-2 infection;
6. To assess how school-children and their families adjust their lives and adopt preventive measures for SARS-CoV-2 over extended periods of time, and how quality of life is affected by the epidemic and preventive measures imposed or recommended by health authorities;
7. To assess how schools adopt preventive measures for SARS-CoV-2 infection over extended periods of time, and how they influence the infection rate;
8. To assess seroprevalence, clustering, and possible routes of transmission to and from children, school personnel and parents.

### Study design

This is a longitudinal, population-based observational study in a regionally-representative cohort of children and adolescents from randomly selected schools and classes in the canton of Zurich, Switzerland. The study is embedded in a Swiss-wide research program *Corona Immunitas* (www.corona-immunitas.ch), where another 25’000 persons (mostly adults) will be enrolled in over 20 prospective studies with fully aligned study protocols.

Children participants were enrolled from June 16^th^ to July 9^th^, 2020, whereas parents and school personnel are enrolled from August 20^th^ to September 5^th^. The follow-up of enrolled children and school personnel is planned until April 2021.

The longitudinal design allows for monitoring the evolution of the epidemic, as well as the impact of school-based and other preventive measures. Three phases are pre-defined, with the possibility for adaptation (i.e., adding Phase IV) according to the dynamic of the pandemic:

- Phase I (June to September 2020): Baseline estimate of seroprevalence of SARS-Cov-2 in school-children, parents and school personnel shortly after the lockdown and subsequent re-opening of schools.
- Phase II (October/November 2020): Estimate of seroprevalence in the same cohort of children and school personnel after the summer holiday and three months of school.
- Phase III (March/April 2021): Estimate of seroprevalence in children and school personnel after the winter season.

### Study setting: primary and secondary schools in Switzerland

One out of six (1.5 million) inhabitants of Switzerland live in the canton of Zurich. The canton is divided into 12 districts (Figure 1). Details on population characteristics and school system are provided in the online data supplement.

**Figure 1.**
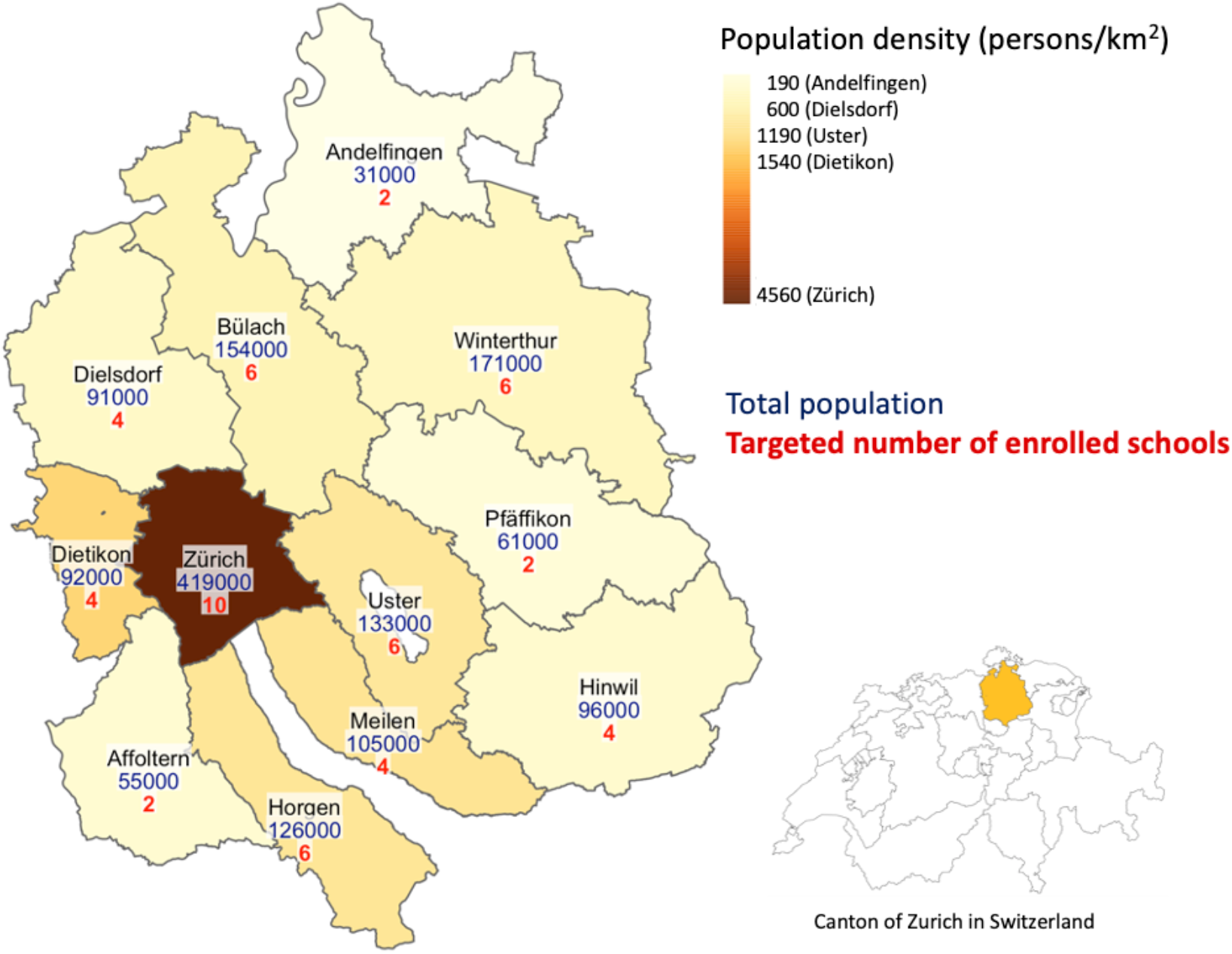
Districts of canton of Zurich: population density, count, and targeted number of enrolled schools

### Ethical aspects

The study was approved by the Cantonal Ethics Committee Zurich (2020-01336). Written informed consent is obtained from parents or legal guardians (referred to as parents further on) of participating children. Children aged 14 years and older can confirm the consent themselves. Additional informed consent is obtained for biobanking plasma samples for subsequent testing within the scope of the SARS-CoV-2 seroprevalence study.

### Schools: sampling and sample size

Random selection of schools is stratified within districts of the canton, and random selection of classes is stratified within lower, middle and upper levels of schools. All children attending the selected classes are invited, except in mixed-age classes (only students from the eligible grades invited).

Primary schools are selected randomly, and the closest secondary school geographically is matched. The targeted number of schools to enroll per district ranges from 2 to 10 depending on the district size. After initial invitation round, school participation rate is assessed and additional schools are selected within required districts, until the aimed number is reached, or further recruitment would not be feasible. Population sizes and targeted number of enrolled schools within districts is depicted in Figure 1.

The overall targeted number of schools is 58 (29 primary and 29 secondary schools). We aim to invite at least 3 classes and at least 40 children per school level. The number of classes and children to invite will be reassessed after calculating the average children participation rate in the first week of enrollment. If needed, additional classes will be invited, aiming to enroll at least 40 children per school level. Assuming a participation rate of 60-80% per class, we would enroll 2100-2800 children in June/July 2020. We expect a seroprevalence rate of 1 to 5% based on the existing research [4]. Depending on the specificity and sensitivity parameters of the test, we expect precision of about ±2%.

### Population: definition

Children and adolescents residing in Switzerland and attending a selected public or private, primary or secondary school (approximate age 5 to 16 years) in the canton of Zurich are eligible for the study, as well as their parents living in the same household, and the entire personnel of participating schools. Main exclusion criteria are small school size (for schools) and suspected or confirmed infection with SARS-CoV-2 during testing (for participants). The 1^st^-2^nd^, 4^th^-5^th^ and 7^th^-8^th^ school grades are included. Third, 6 and 9 grades are excluded as these students may move to another school after the summer break, and follow-up would be compromised. In age-mixed learning classes in primary schools, only 1 and 5 grades are included as children in other grades potentially change the class after the summer break. Detailed inclusion and exclusion criteria are provided in the online data supplement.

## Study procedures

### Recruitment and study timeline

The process of recruitment, communication with invited schools, children and their parents, and testing, is depicted in Figure 2. Randomly selected schools receive an email from the study group, including study information, link to study website (www.ciao-corona.ch), informational videos in multiple languages for schools, parents, and children. Further details on recruitment process are provided in the online data supplement.

**Figure 2.**
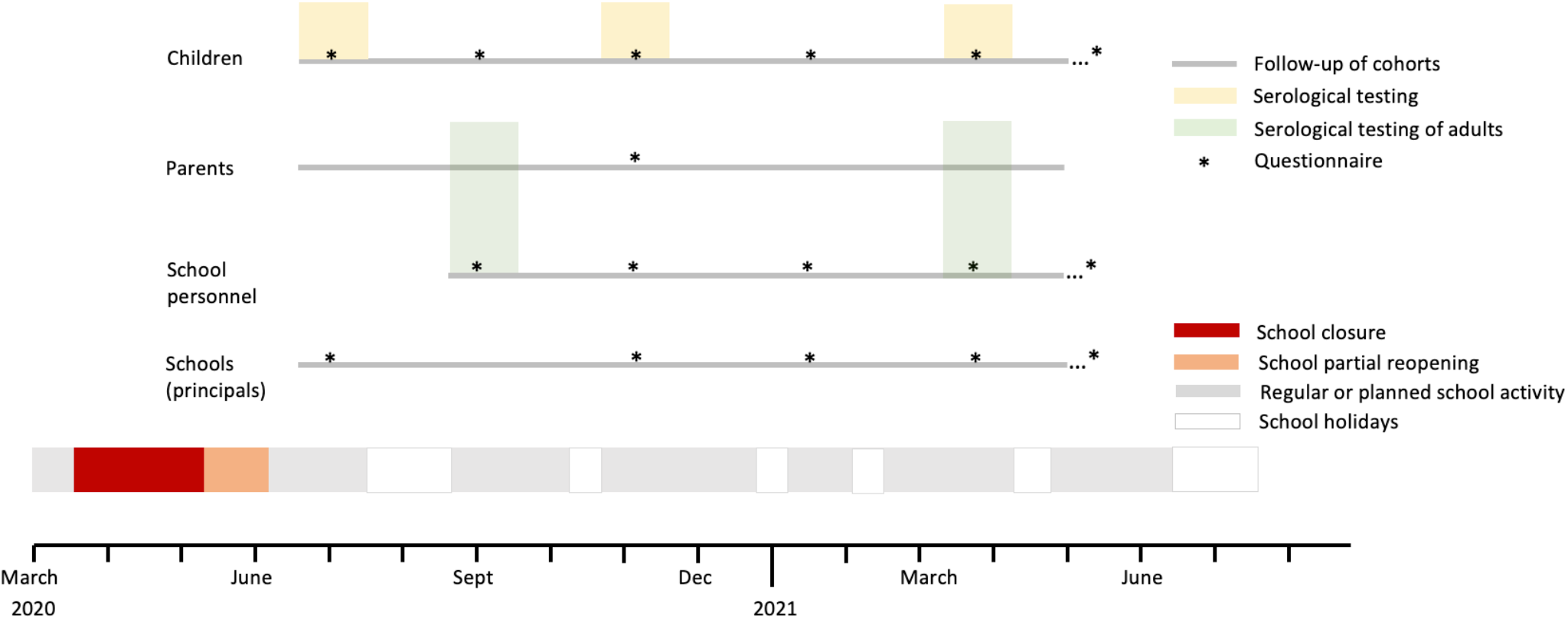
Timeline of study recruitment, serological testing, and follow up Note that the children’s questionnaire includes questions for parents. Timing of testing and questionnaires is approximate and will depend on the development of the pandemic.

### Collection of samples and testing at schools

At each of the planned testing phases, the study team will come to the participating school for half or full day, depending on school size. Testing will take place in a sufficiently large room, in small groups of participating children, with all necessary hygiene and distancing precautions. First, information is provided, child’s identity and consent confirmed, and saliva collected. Venous blood samples will be collected at supine position with the help of anesthetic patches, applied 45-60 min prior to venipuncture. Participating children receive a small age-appropriate gift (worth 5-20$) at the end of each testing.

Adults will be invited for testing in schools or at a testing center. Personal support to fill in the online questionnaire will be offered during testing. Venous blood will be collected. Collected samples are stored, cooled and transported to the laboratory daily after the testing is finished.

### Measurements

Summary information of specimen and questionnaires collected is provided in Table 1.

**Table 1.**
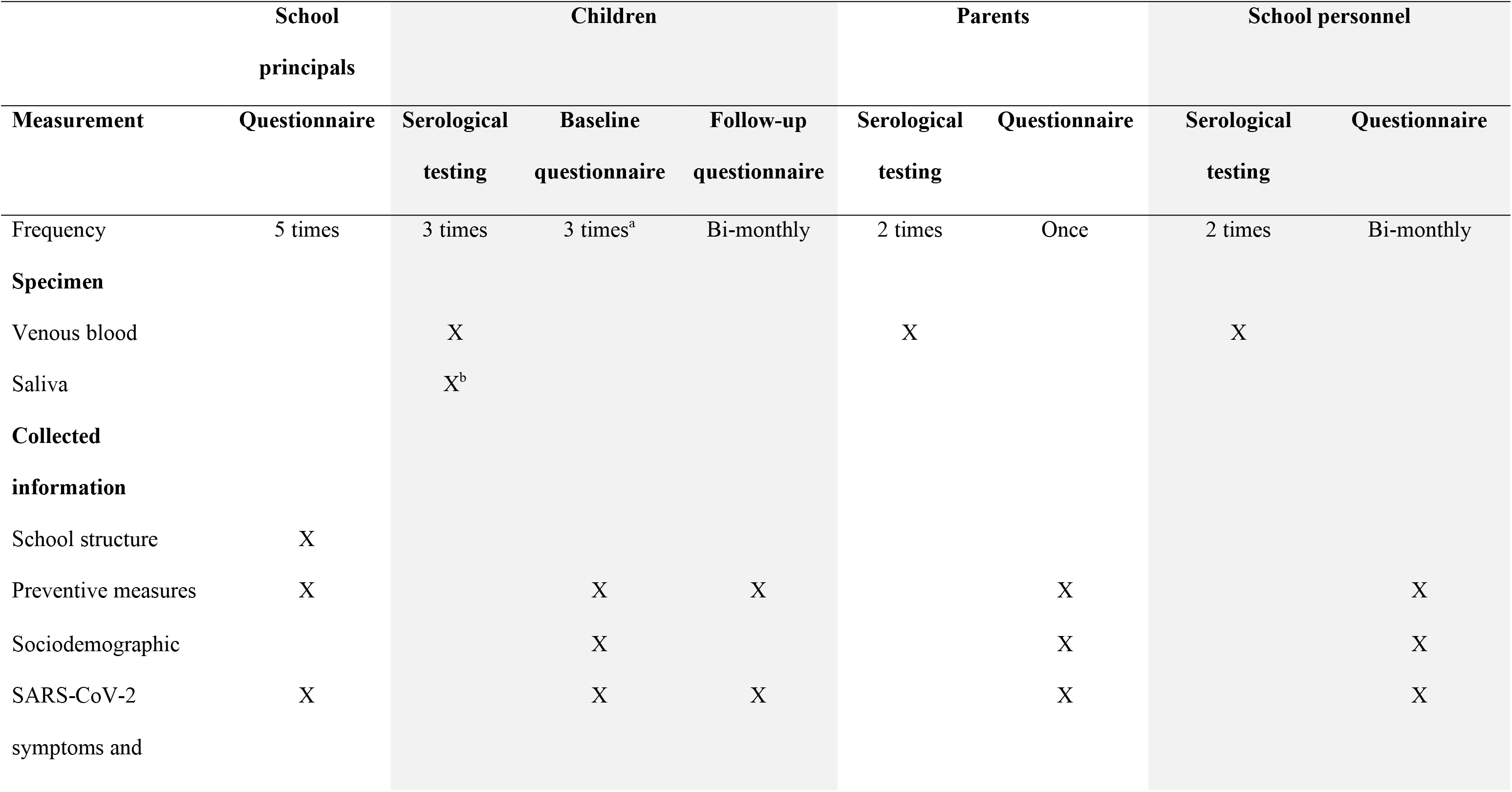
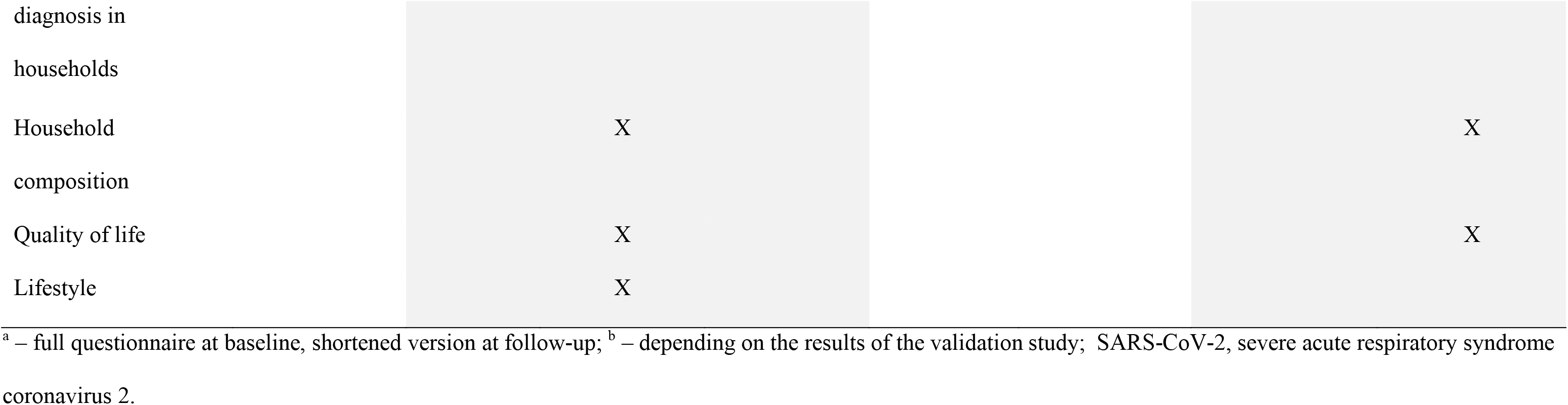
Summary of testing and measurements in study populations

### Blood and saliva samples for serological testing

For each child and adult participant, one sample (9 mL for children and 4.9 mL for adults) of venous blood will be collected for the assessment of SARS-CoV-2 antibodies. Plasma will be separated, aliquoted into 1mL tubes and biobanked at −20°C until testing.

Saliva samples are collected in clean tubes and enriched with virus transport medium. Saliva will first be validated for serological testing. If serological testing in saliva is deemed sufficiently accurate, venous blood sampling might not be necessary in further testing phases. Conversely, if serological testing in saliva is deemed not accurate enough, saliva sample collection will not be continued.

For serological analysis, an in-house developed bead-based binding assay based on the Luminex technology will be used for children. The ABCORA test (version 2.0) provides a highly differentiated picture of the immune response: immunoglobulins G (IgG), M (IgM) and A (IgA) antibodies against four SARS-CoV-2 targets (receptor binding domain (RBD), spike proteins S1 and S2), and the nucleocapsid protein (N) of SARS-CoV-2) are analyzed, resulting in twelve analyzed parameters. Owing to the broad assessment of serological parameters, the ABCORA 2.0 test provides an estimate of infection recency. Based on the ABCORA 2.0 results the seroconversion status of a sample will be classified as positive, weakly reactive, indeterminate, or negative, based on pre-specified threshold values of detected antibody reactivities. In a validation study (unpublished) of 104 samples of SARS-CoV-2 reverse transcription polymerase chain reaction (RT-PCR) positive persons and 251 samples of pre-pandemic, healthy blood donors, test had sensitivity of 93.3-95.2% (compared to sensitivity of 88.5%-93.3% in commercially available tests) and specificity of 98.4-99.6% (depending on the threshold definition of positive and negative cases).

For school personnel and parents, the SenASTrIS (Sensitive Anti-SARS-CoV-2 Spike Trimer Immunoglobulin Serological) assay developed by the Centre Hospitalier Universitaire Vaudois (CHUV), the Swiss Federal Institute of Technology in Lausanne (EPFL) and the Swiss Vaccine Center will be used [16]. The test is used by all study sites of the nationally coordinated research program *Corona Immunitas*.

### Collection of questionnaire data

Detailed information of data collected with each questionnaire is provided in the online data supplement. Briefly, baseline and follow up questionnaires are filled online (when necessary, on paper or over phone) for participating children by parents together with the child, and school personnel (see Figure 2). Follow up questionnaires are sent approximately bi-monthly, adapting to the school year timing, at least until April 2021. School principals will fill in questionnaires at and between each testing phase.

## Study data

### Data management

Study data will be collected in REDCap (Research Electronic Data Capture), a secure, web-based application with access restricted to selected study personnel. The database will also be used to send out online surveys to school personnel and parents, and deliver study results per email. Further details are provided in the online data supplement.

### Data analysis

Descriptive analysis of participant sociodemographic, lifestyle, and behavior information will be performed. Total seroprevalence and cumulative incidence will be calculated, as well as age-, time- and region-specific estimates. In order to also include the sensitivity and specificity of the serological test in the analyses and account for the complex sampling structure (clustering within classes and schools), hierarchical Bayesian logistic regression models will be used.[4] The total numbers of school children in the respective grades per district will be used for post-stratification, so that the estimates are representative for the demographics of the canton of Zurich.

Associations with health and quality of life outcomes will be assessed with multiple regression models. Other planned estimates include proportion of seropositive individuals who have been asymptomatic, risk factors for infection at individual and school level. Associations of levels of IgG, IgM and IgA antibodies with symptoms and risk factors will be assessed.

### Patient and public involvement

The study was initiated together with the Educational Department of the canton of Zurich. Several school principals were consulted during the development of the protocol to ensure feasibility of the planned study procedures. Early feedback was collected from children and parents invited to participate, in order to adapt the communication strategies and channels. Further feedback was collected from enrolled children and school principals during the first testing phase, in order to adapt subsequent testing phases and adult testing. Numerous online informational sessions, encouraging open exchange and feedback, were organized for school principals, personnel and parents of the children. Results of individual tests will be communicated to the participants, and overall study results disseminated to participating schools. Findings will be disseminated in lay language in the national and local press, to the national and regional educational and public health departments and to the website of the study.

## Discussion

This longitudinal population-based cohort study is unique for its focus on children and adolescents in schools. Despite the lacking knowledge how schools contribute to the spread of SARS-CoV-2 infection, major policy decisions on temporary school closure or schedules have been implemented globally in response to the pandemic. This study will contribute to explain the role of schools in order to define the necessary and sufficient preventive measures to balance infection control and impact of school closure.

Currently available population-based seroprevalence studies, which have included children, were mostly conducted in household setting. In May 2020, only 0.8% of children aged 5-9 years were seropositive for SARS-Cov-2 in Geneva, Switzerland, in contrast to 9.6% of children and adolescents aged 10-19 years and 9.9% of adults aged 20-49 years [4]. In April and May 2020, 3.8% of children and adolescents aged 0-19 years were seropositive in Spain, compared to 4.5% to 5.0% in older age groups [5]. In contrast, the current study will primarily consider schools. By analyzing seroprevalence on individual, class and school level, as well as in parents and school personnel, we will be able to identify clusters within these structures. Such knowledge could help to decide if individual classes or whole schools need to be closed to prevent SARS-CoV-2 infection spread. In addition, by testing the entire school personnel, it will inform which employees at schools are the most susceptible to infection.

Only few related planned or ongoing studies have been reported worldwide. In the UK, a study run by Public Health England aims to test seroprevalence in child care facilities and schools in England in May/June, July, and end of autumn 2020 [17]. However, it is not clear if the sampling of schools will be random or stratified by regions or if the structure within schools (classes) will be considered. A smaller study in Berlin, Germany, aims to test 24 randomly selected schools, including 20-40 children and adolescents (500-1000 in total) and 5-10 staff members at each school [18].

The results of this study will likely be generalizable to other cantons in Switzerland and also worldwide, particularly to high- and middle-income countries. The canton of Zurich includes both urban and rural settings, as well as an ethnically and linguistically diverse population. Although the rates of seroprevalence are always location- and time-specific, we believe that the longitudinal design will allow us to investigate many stages of the pandemic.

Due to urgency to launch the study, it faces a few challenges. First, high participation rate of schools and children is required for sufficient power to analyze different regions and clusters within classes and schools. We believe that the high public interest will lead to increased participation, which could otherwise be rather limited in a study collecting venous blood samples in children. In fact, in the initial testing phase in June/July 2020, 55 schools and more than 2500 children were successfully enrolled. Second, the protocol of the longitudinal study will have to be flexible as the pandemic and serological testing methods develop. For this reason, the specific time points of serological testing cannot be fixed in advance (e.g., in case of school closure during the course of the study). Similarly, in further testing phases, it might be sufficient to do the serological testing in saliva or blood, depending on the outcome of the validation of serological testing in saliva. Finally, in order to recruit a sufficient number of schools still before school summer holidays, three rounds of invitations were needed, leading to potential over- or under-sampling of schools in certain districts. However, the sampling discrepancies can be adjusted with weighing of results.

## Conclusions

This population-based cohort study with randomly selected schools and classes across the age range for mandatory school time offers a unique opportunity to observe the longitudinal spread of SARS-CoV-2 infection in children in schools, as well as in their parents and school personnel, thus studying the whole school community. It will report SARS-CoV-2 seroprevalence in children by age groups and regions, provide essential information on possible transmission routes and immunity over time, and assess individual and school-level risk factors for infection. The longitudinal design will allow describing temporal trends of immunity to SARS-CoV-2 and evaluating effects of school structure and preventive measures. It will inform goal-oriented policy decisions in school management during subsequent outbreaks.

## Data Availability

The data for the study, described in this protocol, has not yet been collected.

## Ethics and dissemination

The study was approved by the Ethics Committee of the Canton of Zurich, Switzerland (2020-01336). The results of this study will be published in peer-reviewed journals and will be made available to study participants and participating schools, the Federal Office of Public Health of Switzerland, and the Educational Department of the canton of Zurich.

## Trial registration

ClinicalTrials.gov Identifier: NCT04448717, registered June 26, 2020. https://clinicaltrials.gov/ct2/show/NCT04448717

## Acknowledgements

We greatly appreciate the uncomplicated and supportive collaboration of school principals and teachers, and truly thank all children and their parents for their willingness to participate in this study. We also thank the “Volksschulamt” of the canton of Zurich for providing us with the comprehensive list of all schools and classes of the canton. We would like to thank Jacob Blankenberger (Epidemiology, Biostatistics and Prevention Institute, University of Zurich) for his support in planning the testing of parents and school personnel and Jörg Böni (Institute of Medical Virology, University of Zurich) for his administrative support and expertise in regard to sampling, storage and transport of bio samples.

## Author’s contributions

SK and MP initiated the project and preliminary design, with support of JF. SK, MP, CB, TR, RJ and AU developed the design and methodology. SK, RJ, AU and TR are responsible for the recruitment process and testing at schools. SRH and JB contributed to the design of the statistical methods and statistical analysis plan. Analysis will be performed by SRH and JB. AT and IA contributed to the design of the study methodology and take responsibility for the storage and analysis of samples. AU wrote the first draft of the manuscript. All authors provided edits and critiqued the manuscript for intellectual content.

